# Modelling the outbreak risk of monkeypox virus clade Ib among MSM in the EU/EEA and the impact of targeted vaccination

**DOI:** 10.1101/2025.07.04.25330877

**Authors:** Bastian Prasse, Disa Hansson, Lina Nerlander, Emmanuel Robesyn, Xanthi Andrianou, Carolina Fransson, Anastasia Pharris, Celine Gossner, Helena de Carvalho Gomes, Jose Canevari

## Abstract

Since the upsurge of monkeypox virus (MPXV) clade Ib in the Democratic Republic of the Congo and neighbouring countries in 2024, there have been multiple importations into Europe. So far, secondary transmission from the imported MPXV clade Ib has been limited. However, further importations are likely, and the associated risk of large MPXV clade Ib outbreaks is a public health concern in the EU/EEA. This work assesses the outbreak risk of MPXV clade Ib among men who have sex with men (MSM), since this population group was most affected by previous mpox outbreaks in Europe. Based on a stochastic, individual-based mathematical model on simulated importations and MSM sexual contact network data, our results are twofold. First, if there is an undetected MPXV clade Ib importation into an urban MSM community in the EU/EEA, then we estimate a relatively high probability of around 19% that a small outbreak above 10 cases occurs, and a moderately low probability of around 4% that a large outbreak above 100 cases occurs. Second, we estimate that a pre-emptive increase of the mpox vaccine uptake by 2.5 to 5 percentage points among MSM has a substantial impact on reducing the risk of large outbreaks – this effect is particularly large when targeting the most sexually active individuals. Given continued global circulation of MPXV clade Ib, our work suggests that small-to-moderate-sized outbreaks among MSM in the EU/EEA are likely and that increasing vaccination uptake remains an effective public health measure in reducing the risk of large outbreaks.

## 1 Introduction

The upsurge of MPXV clade Ib cases in the Democratic Republic of the Congo (DRC) and neighbouring countries and limited response capacity prompted the World Health Organization to declare a Public Health Emergency of International Concern in August 2024[1]. Given the continued circulation of MPXV clade Ib outside the EU/EEA and the associated potential for further importations, MPXV clade Ib outbreaks pose a potential public health threat to the EU/EEA. A particular concern is the outbreak risk among men who have sex with men (MSM), since this population has been disproportionally affected by earlier MPXV clade II outbreaks in Europe.

A major global outbreak of mpox occurred in 2022, with over 21,088 cases in the EU/EEA reported to ECDC between May and November 2022 [2]. Cases occurred primarily among MSM [3-5]. A combination of behavioural change, infection-acquired immunity, and public health interventions — including information campaigns, vaccination, contact tracing and case management — effectively reduced viral spread [6,7]. Since then, mpox incidence in the EU/EEA has remained relatively low, with a median number of 122 cases (range from 4 to 232) per month between January 2023 and March 2025 [2].

Two distinct clades of MPXV have been identified based on phylogeny: clade I and clade II, with further subclades Ia, Ib, IIa and IIb. The 2022 global outbreak was caused by MPXV clade II. In contrast, MPXV clades Ia and Ib are the predominant clades circulating in the Democratic Republic of the Congo and neighbouring countries from 1 January 2022 until 25 May 2025, where sustained human-to-human transmission has led to a marked increase in mpox incidence in the DRC in 2023 [8,9]. From August 2024 until May 2025, 21 imported MPXV clade I (1 MPXV clade Ia and 20 MPXV clade Ib) cases without known links to MSM sexual networks have been reported in the EU/EEA, and limited secondary transmission to household or other close contacts has been confirmed [10]. Given the rapid spread of clade II within MSM networks in the EU/EEA in 2022, there is a risk that a future introduction of clade Ib into MSM networks could result in large outbreaks, driven by the heavy-tailed distribution of the number of sexual partners [11].

The risk of an MPXV clade I outbreak in Europe is influenced by the evolving immunity profile of the population, shaped by both historic and recent factors. Following the eradication of smallpox, routine smallpox vaccination — which offers cross-protection against mpox — was discontinued in many countries between 1970 and 1984, leaving younger cohorts largely unprotected [12,13]. The 2022 mpox outbreak led to vaccination campaigns in the EU, primarily targeting MSM, with varying levels of uptake across countries in the EU/EEA [14,15]. There is currently no evidence suggesting that vaccine effectiveness against infection differs between clade I and clade II, which is estimated at around 82% [16]. Natural immunity acquired by those infected during the 2022 outbreak further contributes to the current immunity landscape [6,17].

The outbreak of MPXV clade Ib among MSM in the EU/EEA requires two successive events. First, MPXV clade Ib is imported into the EU/EEA by an undetected infectious man who has sex with men, who acquired the infection in a country with ongoing clade Ib circulation. It is challenging to accurately estimate the probability of this first event ^1^ due to various uncertain factors, both within and outside of Europe. However, provided continued global circulation of MPXV clade Ib, an importation into MSM networks seems increasingly probable. Second, the infectious individual causes an MPXV clade Ib outbreak via sexual contacts in an MSM network, where the key public health concern is the potential for causing a large outbreak.

In this work, we use mathematical modelling to estimate the risk of MPXV clade Ib outbreaks among MSM communities in the EU/EEA (the second event) *given that* an undetected importation (the first event) has occurred. Furthermore, we assess the benefit of campaigns of increasing the mpox vaccination uptake among MSM communities. Our results aim to support public health professionals in the EU/EEA in anticipating, preparing and responding, especially by vaccination campaigns, to potential future MPXV clade Ib outbreaks.

## 2 Methods

We developed an individual-based stochastic SEIRV (susceptible–exposed–infectious–recovered–vaccinated) model to simulate the spread of MPXV among MSM in the EU/EEA, see Supplement S2 for more details. The model runs on a sexual contact network and does not consider reinfections. Individuals who recovered from prior MPXV clade II infection were assumed to be immune to clade Ib. The simulated population consists of N = 10,000 individuals — broadly reflecting the size of the MSM population in a relatively large EU/EEA city^2^. The sexual contact patterns are described by a configuration network model with two layers: one for steady sexual partnerships and one for non-steady sexual contacts, both parameterised using data from EMIS-2017 [20]. Model parameters for MPXV transmissibility and naturally acquired immunity were calibrated using case data reported to TESSy from 2022-2024, accounting for underreporting. Specifically, we considered detection probabilities of 40% and 90% to represent worst-case and best-case scenarios, respectively. We simulated the spread of MPXV clade Ib among MSM starting with a single undetected infectious individual, which is selected at random with a probability proportional to the individual’s total number of sexual contacts to reflect differences in potential exposure risk^3^. For each parameter setting, we ran the stochastic model 1,000 times.

In addition to natural immunity, the model includes vaccine-induced protection from two sources: historical smallpox vaccination and mpox-specific vaccination campaigns initiated during the 2022 outbreak. Based on published estimates, we assume that 14.8% of the MSM population are protected against MPXV infection due to historical smallpox vaccination, after accounting for waning of immunity [13].

To estimate the mpox-specific vaccine uptake among MSM^4^, we obtain the denominator by estimating the number of MSM in each EU/EEA country by assuming that 50% of the population is male, 2.8% of men are MSM, and 67% of MSM are sexually active (67% approximately equals the proportion of men in the EU/EEA aged 16-65 years). Since full vaccination requires two doses, we divide the reported number of doses administered until September 2024 (data collected by ECDC, unpublished) by two to obtain an estimate of the number of fully vaccinated individuals. The resulting proportion of fully vaccinated MSM ranges from 0%-13.18% across countries in the EU/EEA with a median of 2.5%. (50% UI: 0.36-8.29%). To capture this variation across MSM communities in the EU/EEA, we varied vaccine coverage between 0% and 10% in our simulations. We assumed a vaccine effectiveness against infection of 82% [16] and no reduced infectivity of vaccinated individual with a breakthrough infection. Furthermore, we consider preferential vaccine allocation to individuals with higher sexual activity. Specifically, we explore two scenarios in which vaccines were distributed randomly to either the top 15% or 30% of individuals with the most sexual partners. Booster doses were not considered.

We modelled contact tracing by classifying each infected individual into one of three mutually exclusive categories: *primary detected, secondary detected*, and *undetected*. The first category, primary detected individuals, are those that develop symptoms, seek care, and are diagnosed, thereby triggering contact tracing with a certain probability. This probability reflects potential limitations in public health resources and individual willingness to disclose contacts. Before detection, primary detected (or rather “to-be primary detected”) individuals can infect others, which includes presymptomatic transmission. Upon detection, primary detected individuals are assumed to self-isolate. The second category, secondary detected individuals, are sexual contacts identified through contact tracing of a primary case. Before being traced, secondary detected individuals can infect others. Once traced, secondary detected individuals self-isolate on the same day, and onward tracing of their contacts is initiated. We assumed that steady sexual partners are more likely to be successfully traced than casual contacts. The third category, undetected individuals, are neither identified through symptoms nor via contact tracing. Given uncertainty in detection and contact tracing probabilities, as well as variations of these probabilities across MSM communities in the EU/EEA, we conducted simulations across a range of parameter values (see Supplementary Table S1).

The increased circulation of MPXV clade Ib starting in August 2024, raises concern that MPXV clade Ib is more transmissible than MPXV clade II. However, there is limited evidence on the level of transmissibility of MPXV clade Ib relative to clade II. Hence, to account for this uncertainty regarding the transmissibility, we considered two scenarios in this work: A baseline transmissibility scenario^5^ assuming equal transmissibility, and an increased transmissibility scenario assuming that clade Ib is 15% more transmissible than clade II.

## 3 Results

### 3.1 The probability of an MPXV clade Ib outbreak among MSM following the importation of a single case

Figure 1 shows the estimated probability of an MPXV clade Ib outbreak starting from a single infectious individual, for both the baseline and increased transmissibility scenarios. For the precise numbers underlying Figure 1, we refer to Supplementary Table S4. We highlight three observations:

**Figure 1.**
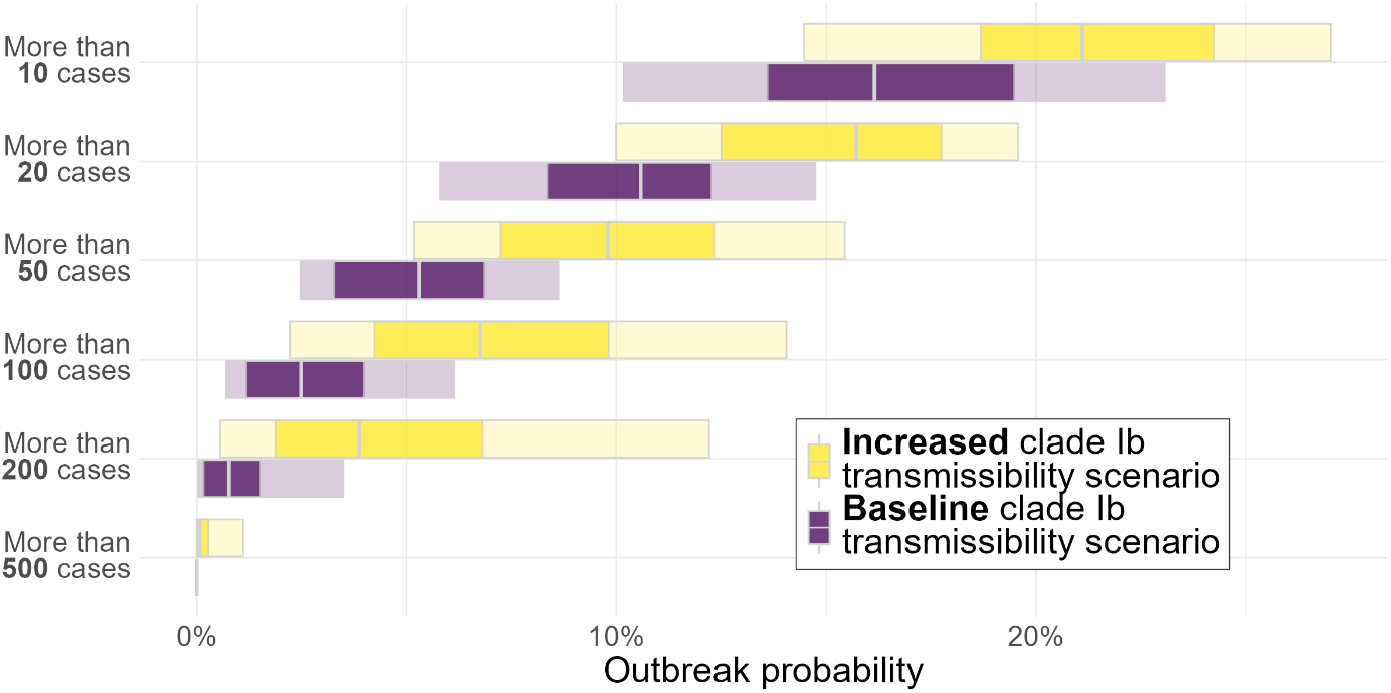
Probability of an MPXV clade Ib outbreak of varying sizes in an MSM population starting with a single undetected infectious individual. Results are shown for different cumulative incidence thresholds under two transmissibility scenarios: a baseline scenario assuming clade Ib is equally transmissible as clade II, and an increased transmissibility scenario assuming clade Ib is 15% more transmissible. Bars represent uncertainty due to different characteristics of MSM populations across cities in the EU/EEA, including variation in sexual contact patterns and immunity levels due to mpox vaccine uptake and prior clade II infections. Dark and light bars show the 50% and 90% uncertainty intervals (UI), respectively; the grey line indicates the median.

- **Outbreaks exceeding 10 cases occur with a relatively high probability** of 15.0% (90% UI: 9.6-21.7%) and 20.3% (90% UI: 13.7-25.6%) for the baseline and increased transmissibility scenario, respectively. Combining both transmissibility scenarios yields an outbreak probability of 19.1% (90% UI: 11.2-26.4%).
- **Large outbreaks are substantially less likely than small outbreaks.** For instance, outbreaks over 200 cases occur with a probability of 0.7% (90% UI: 0.0-3.5%) and 3.9% (90% UI: 0.6-12.2%) for the baseline and increased transmissibility scenario, respectively.
- **In the increased transmissibility scenario, the probability of large outbreaks is markedly higher, while smaller outbreaks are less affected.** For example, the probability of outbreaks exceeding 200 cases is about 450% more likely under the higher transmissibility scenario as compared to the baseline transmissibility scenario (comparing the median probabilities 3.9% vs. 0.7%). In contrast, the probability of outbreaks exceeding 10 cases is only about 35% more likely (20.3% vs. 15.0%).

### 3.2 The impact of increasing the mpox vaccination uptake

We estimated the impact of increasing the mpox vaccination uptake among MSM on reducing the probability of a clade Ib outbreak. The results are presented by combining the baseline and the higher transmissibility scenario. The additional vaccine rollout builds on the uptake as of November 2024. Three vaccine uptake scenarios were considered: 1) a **baseline scenario with no additional vaccine uptake**, 2) a scenario with a **low vaccine uptake increase of 2.5 percentage points**, and 3) a scenario with a **high vaccine uptake increase of 5 percentage points**.

For each uptake scenario, we evaluated three levels of vaccine targeting, based on sexual activity. In the **high**-**targeting scenario**, vaccines are distributed randomly among the **15% of MSM with the most sexual partners**. The **medium- and low-targeting scenarios** assumed vaccine distribution among the **25% and 50% most sexually active individuals**, respectively. These targeting scenarios are not intended to define practical implementation strategies, but rather to provide qualitative insights into the potential added benefit of targeted vaccination campaigns.

In summary, a total of six increased vaccination scenarios (three targeting levels times two uptake levels) and one baseline scenario with no increased uptake were analysed, as shown by Table 1. We emphasise that not everyone in the target group is vaccinated, since the modelled increased vaccine uptake (of either additional 2.5% or 5% of the total MSM population) is smaller than the target group sizes.

**Table 1.**
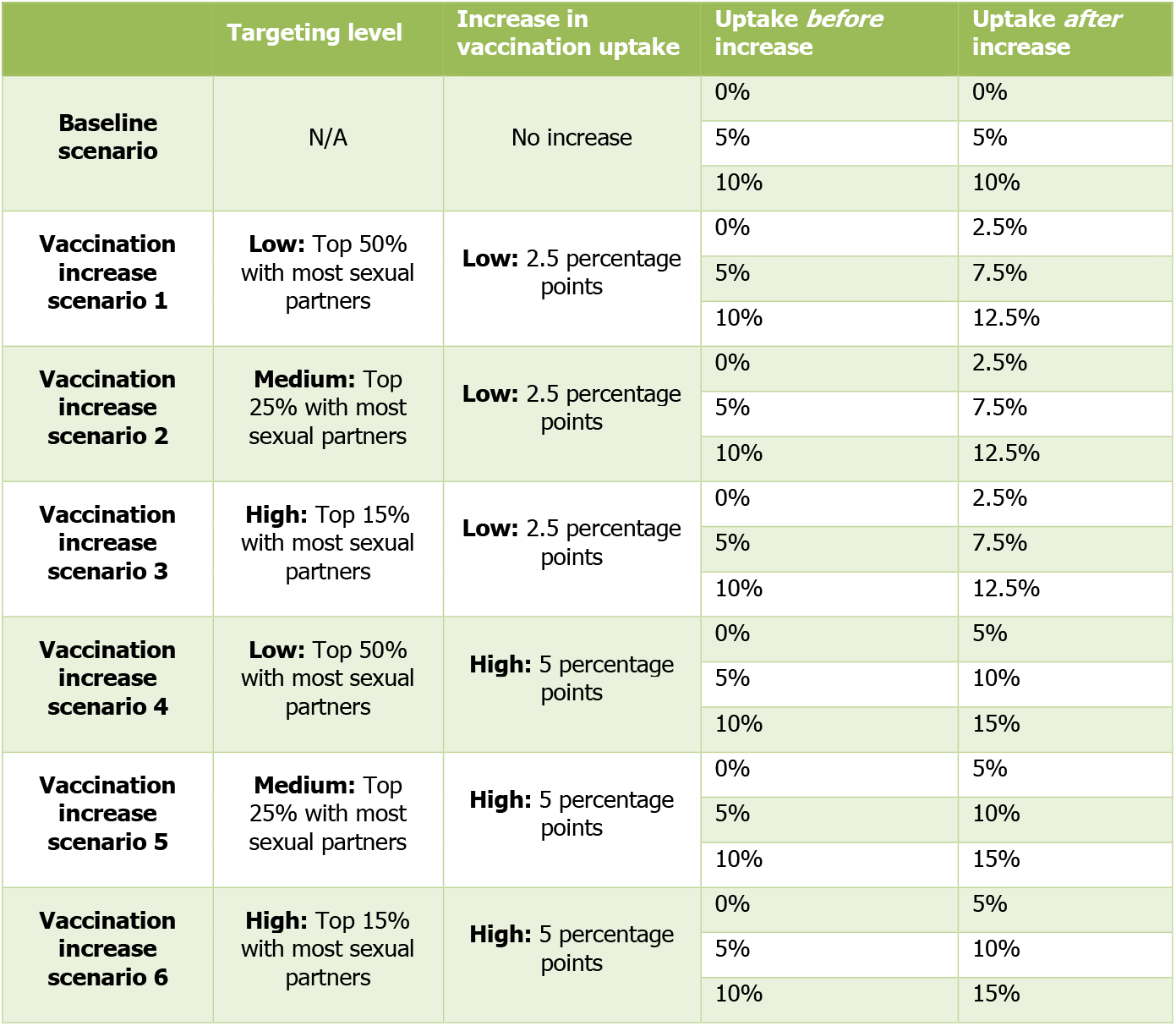
Overview of the baseline scenario and six vaccination increase scenarios evaluated in this analysis. Scenarios vary by level of additional vaccine uptake among MSM (0, +2.5, or +5 percentage points) and the degree of targeting based on sexual activity (top 15%, 25%, or 50% of MSM with the most sexual partners). Note that, since the initial vaccination uptake among MSM before the increase is unknown, we explore three different initial uptakes (0%, 5%, 10%). These three initial uptakes form sub-scenarios for each of the six increase scenarios and the baseline scenario. Therefore, a total of 21 scenarios (7 increase/baseline scenarios times 3 sub-scenarios) is considered.

|  | Targeting level | Increase in vaccination uptake | Uptake *before* increase | Uptake *after* increase |
| --- | --- | --- | --- | --- |
| **Baseline scenario** | N/A | No increase | 0% | 0% |
|  |  |  | 5% | 5% |
|  |  |  | 10% | 10% |
| **Vaccination increase scenario 1** | **Low:** Top 50% with most sexual partners | **Low:** 2.5 percentage points | 0% | 2.5% |
|  |  |  | 5% | 7.5% |
|  |  |  | 10% | 12.5% |
| **Vaccination increase scenario 2** | **Medium:** Top 25% with most sexual partners | **Low:** 2.5 percentage points | 0% | 2.5% |
|  |  |  | 5% | 7.5% |
|  |  |  | 10% | 12.5% |
| **Vaccination increase scenario 3** | **High:** Top 15% with most sexual partners | **Low:** 2.5 percentage points | 0% | 2.5% |
|  |  |  | 5% | 7.5% |
|  |  |  | 10% | 12.5% |
| **Vaccination increase scenario 4** | **Low:** Top 50% with most sexual partners | **High:** 5 percentage points | 0% | 5% |
|  |  |  | 5% | 10% |
|  |  |  | 10% | 15% |
| **Vaccination increase scenario 5** | **Medium:** Top 25% with most sexual partners | **High:** 5 percentage points | 0% | 5% |
|  |  |  | 5% | 10% |
|  |  |  | 10% | 15% |
| **Vaccination increase scenario 6** | **High:** Top 15% with most sexual partners | **High:** 5 percentage points | 0% | 5% |
|  |  |  | 5% | 10% |
|  |  |  | 10% | 15% |

Figure 2 shows the probability of a clade Ib outbreak in the baseline scenario and the estimated reduction in outbreak probability under the six increased vaccination scenarios following the importation of MPXV clade Ib into the synthetic MSM population. (For each of the six vaccination increase scenarios and the baseline scenario, the results of the three respective sub-scenarios are combined.) For the precise numbers underlying Figure 2, please see Supplementary Table S5. Four key observations emerge:

**Figure 2.**
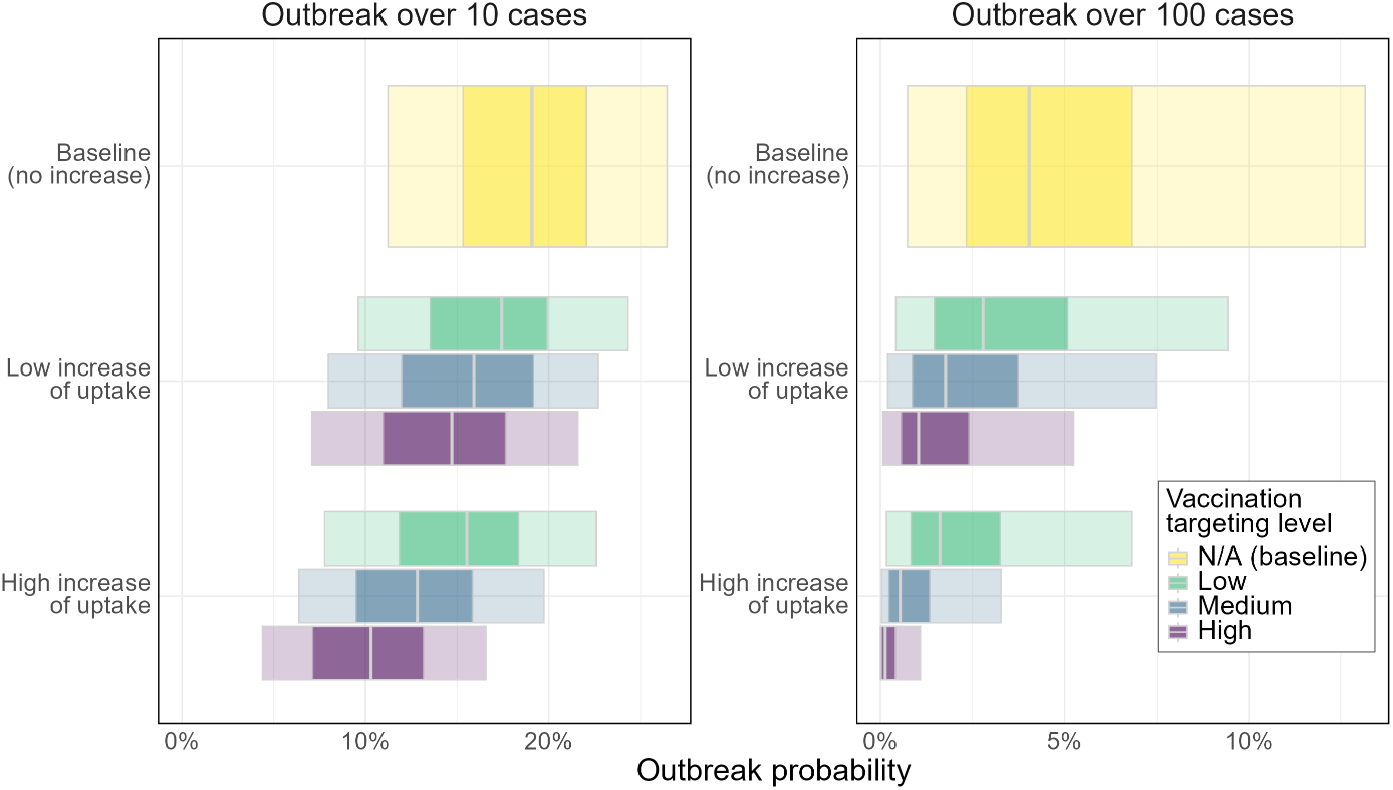
Effect of increased vaccine uptake on the probability of an MPXV clade Ib outbreak exceeding 10 or 100 cases in an MSM population, following the importation of MPXV clade Ib by a single undetected infectious individual. Note the differing x-axis scale across subplots. Bars represent different combination of vaccine uptake and targeting strategies. Each bar captures uncertainty due to different characteristics of MSM populations across cities in the EU/EEA, including variations in sexual contact patterns and immunity levels from current mpox vaccine uptake (as of March 2025) and past clade II infections. Results combine both clade Ib transmissibility scenarios. Dark and light bars indicate the 50% and 90% uncertainty intervals, respectively; the grey line shows the median.

- **Even a modest increase in vaccine uptake can substantially reduce outbreak risk.** An increase of only 500 doses (i.e., given 2 doses per full vaccination, 250 fully vaccinated individuals among the 10,000 MSM) reduced the median probability of outbreaks exceeding 100 cases by 30% under low targeting level (4.0% vs 2.8%), and by 75% under high targeting level (4.0% vs 1.0%).
- **Vaccination is particularly effective in preventing large outbreaks.** For example, under a high uptake increase and medium targeting level, the probability of an outbreak exceeding 10 cases declined by ~33% compared to the baseline (19.1% vs 12.8%), while the probability of an outbreak exceeding 100 cases is reduced by ~88% (4.0% vs 0.5%).
- **Targeting vaccination to sexually active individuals is critical for effectiveness.** To prevent outbreaks, a low uptake increase with medium to high targeting is more effective than a high uptake increase with low targeting, highlighting the importance of prioritisation strategies.
- **Large outbreaks become highly unlikely when moderate or high targeting is achieved.** For example, with a medium targeting level, the probability of outbreaks exceeding 100 cases dropped to 0.5% (90% UI: 0.0-3.3%) and 0.1% (90% UI: 0.0-1.1%) for a low and high uptake increase, respectively.

## 4 Discussion and conclusions

The main findings of this study are threefold. First, we estimate the probability of a small outbreak (above 10 cases) to be relatively high, while the probability of a large outbreak (above 100 cases) is estimated to be moderately low – these probabilities are *conditional on* a single undetected importation of MPXV clade Ib into an MSM community in the EU/EEA. Even though the probability for a large outbreak is low, provided ongoing MPXV clade Ib circulation outside the EU/EEA, the risk of *multiple* undetected importations rises — and with it, the probability of a large outbreak (see Supplementary Figure S2).

Second, we showed that the transmissibility of MPXV clade Ib is a crucial factor influencing the risk of large outbreaks. However, it is unclear if, and to what extent, MPXV clade Ib is more transmissible than clade II. Gathering further evidence on the transmissibility is essential for more accurate estimates of the potential of large MPXV clade Ib outbreaks among MSM and among the general population.

Third, on agreement with other work [21-23] on vaccination in response to the MPXV clade II outbreak, our model shows that pre-exposure vaccination, particularly among high-risk MSM, remains a highly effective strategy for preventing and containing MPXV clade Ib outbreaks. Notably, even a modest increase in vaccine uptake — by just 2.5 percentage points among sexually active MSM — led to a marked reduction in the probability of small outbreaks and rendered large outbreaks highly unlikely.

Although this analysis focusses on MPXV clade Ib, the results might also hold for MPXV clade Ia – provided similar transmissibility of clade Ia and clade Ib. Furthermore, our work could inform the risk of MPXV clade II outbreaks, if one focusses only on the baseline scenario which assumes that clade Ib and clade II have the same transmissibility. Specifically, the modelling framework could be used as a basis for evaluating the level of vaccination or behavioural intervention required to reduce the risk of ongoing clade II outbreaks in the EU/EEA to very low levels.

A key strength of our mpox modelling approach is the incorporation of a contact network structure that captures heterogeneity in sexual activity levels within the MSM population. These activity levels were informed by the EMIS-17 survey [20]. We acknowledge, however, that sexual behaviours and partnership patterns may have evolved since 2017, potentially influenced by changes in sexual health practices — such as the increased uptake of HIV pre-exposure prophylaxis (PrEP) — which has been associated with changes in condom use and partner dynamics in some MSM populations [24,25]. Updating the model with data from future iterations of the EMIS survey (EMIS has been repeated in 2024, and an update is ongoing) or other recent behavioural studies would likely enhance its accuracy and relevance. Another strength of the model is its ability to account for several important uncertainties, including current vaccine coverage, the extent of naturally acquired immunity, and the effectiveness of contact tracing.

While this study emphasises sexual transmission of MPXV among MSM, other modes of transmission — such as heterosexual and household transmission — are also possible. Household transmission following importation of clade Ib cases has been reported within the EU/EEA, namely in Germany and Belgium [10]. However, based on modelling evidence from the U.S. CDC [26], household transmission is considered less likely to result in large-scale outbreaks in the EU/EEA compared to sexual transmission among MSM.

This study has several limitations and is based on key assumptions. We emphasise that this study only considers the outbreak risk among MSM given an undetected importation, hence, we do not cover the probability that MPXV clade Ib is imported and not detected. In the absence of data on cross-immunity between MPXV clades, we assumed complete cross-immunity.

Additionally, post-exposure vaccination of contact-traced individuals was not included in the model. The Markovian modelling approach used in this analysis assumes an exponential distribution for detection and tracing delays; however, a right-skewed distribution may be more appropriate. Given the limited availability of data on contact tracing parameters, we explored a range of plausible values (see Supplementary Table S1). More accurate parameter estimates would improve the precision of our findings.

## Data Availability

Access to data can be requested from the European Centre for Disease Prevention and Control.

## Acknowledgements

We thank Nathalie Nicolay and Sabrina Bacci for contributing to this work. Furthermore, we are grateful for the valuable input by Vítor Cabral Veríssimo, Mariana Perez Saad Duque, Constantino Pereira Caetano, Asunción Díaz Franco, David Garcia-Garcia, Derval Igoe, and the Swedish mpox outbreak response team for their expert opinion guiding the scenario design for the contact tracing parameters.

## Supplementary material

### S1. Disease parameters

Supplementary Table S1 summarises the parameters of this work. For further details on the function of these parameters, we refer to Section S2.

**Supplementary Table S1.**
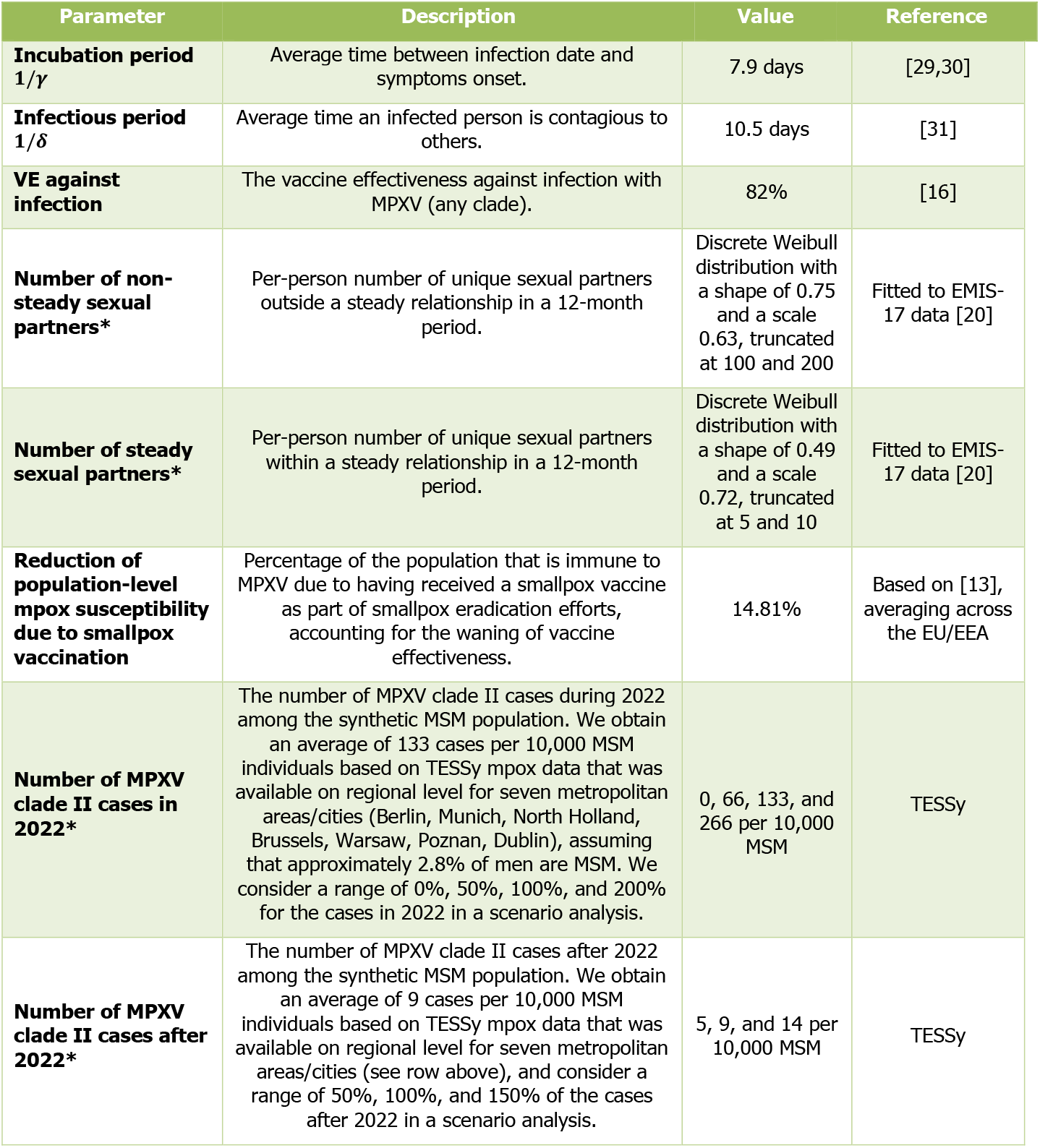
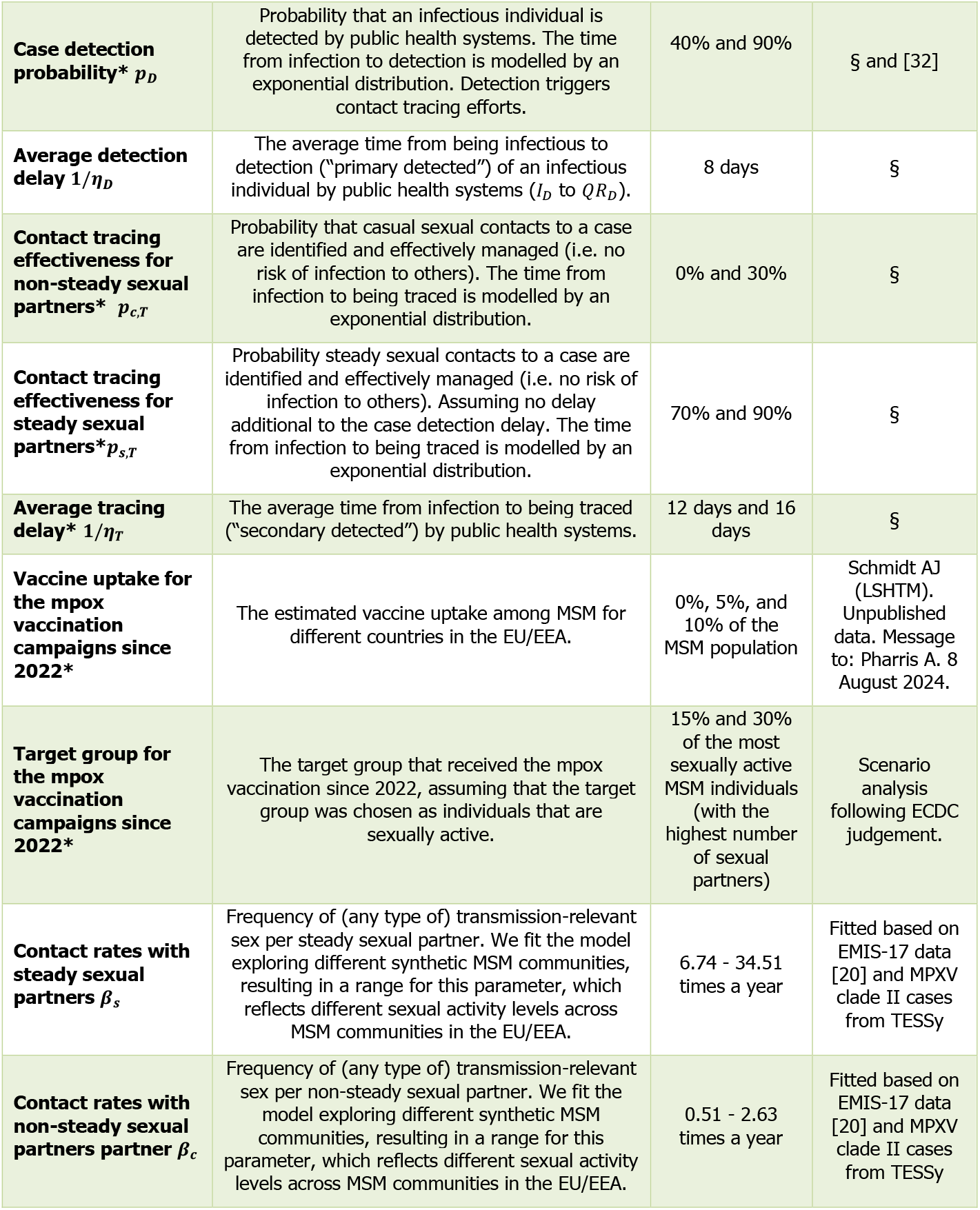
Description of parameters used in this modelling study. To account for parameter uncertainty, simulations were run varying scenarios for the parameters marked with a star (*), where the four contact tracing parameters p_D_, p_T_, η_D_, η_T_ were varied all at once. In total, this resulted in 4×3×2×3=72 different parameter sets, which capture the heterogeneity of MSM communities across the EU/EEA. The results of the 72 parameter sets were then combined to produce an aggregate model outcome. (§) Expert assessment following personal communication with Vítor Cabral Veríssimo, Mariana Perez Saad Duque, Constantino Pereira Caetano, Asunción Díaz Franco, David Garcia-Garcia, Derval Igoe, and the Swedish mpox outbreak response team and ECDC staff, and informed by [27,28].

### S2. Details of the mathematical model

#### S2.1 Compartments and transitions

**Supplementary Figure S1.**
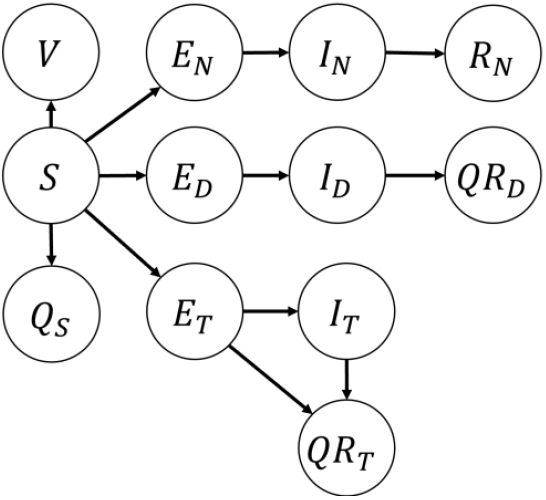
Disease states (compartments) and transitions of the mathematical model. Note that the viral spread among the population is modelled on individual level via a sexual contact network and, therefore, individuals with the same disease state are not interchangeable.

Supplementary Figure S1 provides an illustration of the disease states of the individual-based model and the transitions between the states. As explained in the main text, we model contact tracing by assigning individuals into either one of three categories, each of which corresponds to a set of compartments:

‐ Individuals that are (will become) primary detected are in either of the compartments *E_D_, I_D_, QR_D_*. This only includes individuals that adhere to self-isolation upon primary detection, and the fraction of primary detected individuals that do not comply are moved in the ‘undetected’ track (*E_N_, I_N_, R_N_*).
‐ Individuals that are (will become) secondary detected (“traced”) are in either of the compartments *E_T_, I_T_, QR_T_*. This only includes individuals that adhere to self-isolation upon secondary detection, the fraction of secondary detected individuals that do not comply are moved in the ‘undetected’ track (*E_N_, I_N_, R_N_*).
‐ Undetected individuals are in either of the compartments *E_N_, I_N_*, or *R_N_*.

The individual-based mathematical model describes the disease progression by 12 compartments:

‐ Individuals in the **susceptible *S*** compartment are susceptible to infection, which includes unvaccinated individuals, unsuccessfully vaccinated individuals (due to a vaccine effectiveness smaller than 100%), and individuals who were instructed to quarantine but do not comply.
‐ Individuals in the **vaccinated *V*** are successfully vaccinated and cannot become infected.
‐ Individuals in the **quarantined susceptible *Q_S_*** are quarantined following a contact with an infectious individual which did not result in an infection.
‐ Individuals in the **exposed *E_N_, E_D_, E_T_*** compartments have acquired the disease but are not infectious yet. The distinction based on the three subscripts is explained above. Note that the three exposed compartments are identical except for which compartments they transition towards, which eventually terminates in either *R_N_, QR_D_*, or *QR_T_*.
‐ Individuals in the **infectious *I_N_, I_D_, I_T_*** compartments are infectious. The distinction based on the three subscripts is explained above.
‐ Individuals in the **recovered *R_N_*** compartment are recovered from the disease and are undetected. Recovered individuals are immune to reinfection.
‐ Individuals in the **isolated and/or recovered *QR_D_* and *QR_T_*** compartments are in self-isolation and/or recovered from the disease, where the former compartment corresponds to primary detected and the latter to secondary detected individuals.

We emphasise that, for computational efficiency, the model is based on a simplified implementation of contact tracing. In particular, the model compartments do not keep track of the exact history of who-infected-whom. For instance, two individuals that are traced infectious are both in the compartment *I_T_*, even if they acquired the infection from different infectors. As a consequence, this simplification does not fully capture the sequence of contact trancing. For instance, suppose that a to-be-detected infector *j* in *I_D_*(who is detected once transitioning to *QR_D_*) infects individual *i*, who transitions into the to-be-traced, exposed compartment *E_T_*. Since the transitions follow a stochastic process, it can occur that – following a tracing event – the infected individual *i* transitions into the compartment *QR_T_ before* the infector *j* is detected, i.e., before the transition of individual *j* from compartment *I_D_* to *QR_D_*. In such cases, the sequence of contact tracing is not correct. Instead of accurately describing the sequence of contact tracing events, the model aims to capture the temporal average effect of contact tracing.

We define the **incubation period 1/*γ***, the **infectious period 1/*δ***, the **detection delay 1/*η_D_***, and the **tracing delay 1/*η_T_***, see Supplementary Table S1 for a description of these parameters. Then, we obtain the transition between the compartments as specified by Supplementary Table S2.

**Supplementary Table S2.**
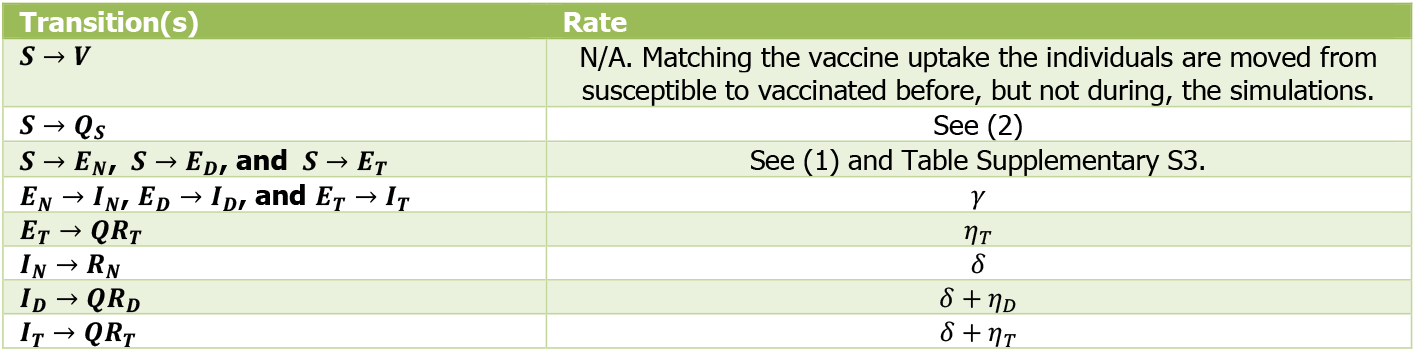
Transition rates of the mathematical model. Since the transitions from S to Q_S_ and the exposed compartments are more complicated, they are defined below this table. Note that the transition from I_D_ to QR_D_ occurs through recovery (with rate δ) or through self-isolation (with rate η_D_), which results in the total rate δ + η_D_, and similarly for the transition from I_T_ to QR_T_.

The transition rate from the susceptible compartment to the exposed compartments *E_N_, E_D_, E_T_* is more complicated and depends on the number of infectious individuals in *I_N_, I_D_, I_T_*. We denote the rate of a susceptible individual *i* to become infected at time *t*, i.e., to transition from to either of the three exposed compartments, as

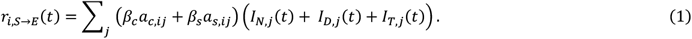

Here, we sum over all individuals *j* in the MSM population. If individuals *i* and *j* are connected via a non-steady (casual) sexual partnership, then *a*_*c*,*ij*_ = 1; otherwise, *a*_*c*,*ij*_ = 0. Analogously, *a*_*s*,*ij*_ ∈ {0,1} indicates whether individuals *i* and *j* are connected via a steady sexual partnership. The contact rates on the non-steady and steady layer, *β_c_*and *β_s_*, are obtained from the model calibration (see Section S2.5). If individual *j* is in the compartment *I_N_* at time *t*, then *I*_*N*,*j*_(*t*) = 1; otherwise, *I*_*N*,*j*_(*t*) = 0. Similarly, the functions *I*_*D*,*j*_(*t*) and *I*_*T*,*j*_(*t*) indicate whether individual *j* is in compartment *I_D_* and *I_T_* at time *t*, respectively.

Given that a susceptible individual *i* becomes infected at time *t* (exponentially distributed with rate r_*i*,*S*→*E*_(*t*)), the conditional probabilities that individual *i* transitions to compartment *E_N_, E_D_*, and *E_T_* depend on the source of the infection (the infector) and are specified by Supplementary Table S3. Here, in agreement with Supplementary Table S1, we define the case detection probability *p_D_*, the contact tracing effectiveness for non-steady sexual partners *p*_*c*,*T*_, and the contact tracing effectiveness for steady sexual partners *p*_*s*,*T*_.

**Supplementary Table S3.**
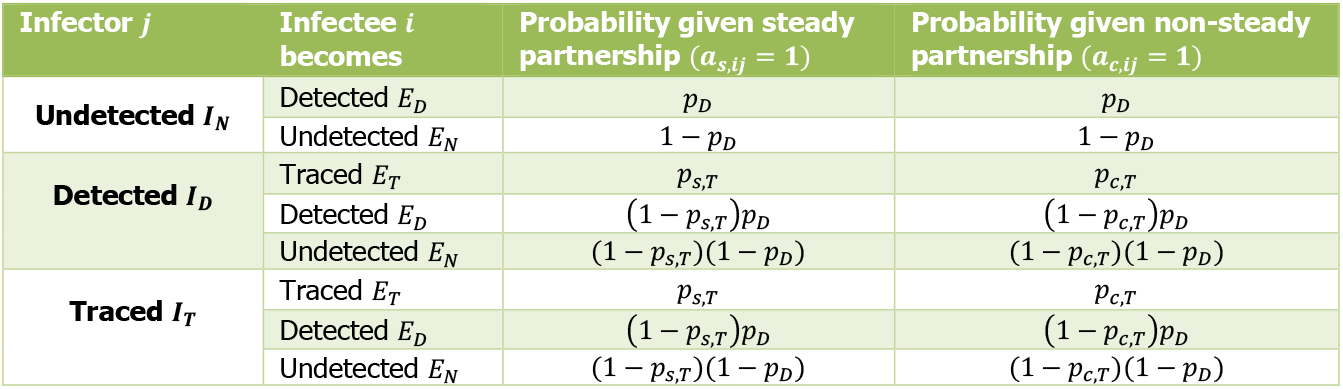
Conditional probabilities of a susceptible individual i transitioning to either of the exposed compartments E_N_, E_D_, E_T_, given that individual i becomes infected at time t. The conditional transition probabilities depend on the state of the infector j from whom individual i acquired the infection.

| Infector $j$ | Infected $i$ becomes | Probability given steady partnership ($a_{s,ij} = 1$) | Probability given non-steady partnership ($a_{c,ij} = 1$) |
| --- | --- | --- | --- |
| **Undetected $I_N$** | Detected $E_D$ | $p_D$ | $p_D$ |
| | Undetected $E_N$ | $1 - p_D$ | $1 - p_D$ |
| **Detected $I_D$** | Traced $E_T$ | $p_{s,T}$ | $p_{c,T}$ |
| | Detected $E_D$ | $(1 - p_{s,T})p_D$ | $(1 - p_{c,T})p_D$ |
| | Undetected $E_N$ | $(1 - p_{s,T})(1 - p_D)$ | $(1 - p_{c,T})(1 - p_D)$ |
| **Traced $I_T$** | Traced $E_T$ | $p_{s,T}$ | $p_{c,T}$ |
| | Detected $E_D$ | $(1 - p_{s,T})p_D$ | $(1 - p_{c,T})p_D$ |
| | Undetected $E_N$ | $(1 - p_{s,T})(1 - p_D)$ | $(1 - p_{c,T})(1 - p_D)$ |

Lastly, the transition for an individual *i* in the susceptibly compartment *S* to the quarantined compartment *Q_S_* at time *t* follows an exponential distribution with rate

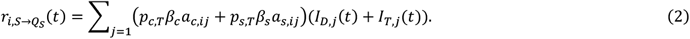

Here, only infected individuals that are either primary detected (*I*_*D,j*_(*t*) = 0) or secondary detected/traced (*I*_*T,j*_(*t*) = 0) contribute to the transition rate from compartment *S* to *Q_S_*.

#### S2.2 Sexual contact network

We describe the sexual contact patterns among MSM by a two-layered configuration model, which is informed by EMIS-17. To obtain the degree distribution of the non-steady and steady sexual contacts in the MSM network, we rely on the data reported by Table 5.7 (“Numbers of steady partners in the last 12 months among men who had ever had sex with a man”) and Table 5.8 (“Numbers of non-steady partners in the last 12 months among men who had ever had sex with a man”) in the EMIS-17 report. In particular, to capture the heavy tail of the degree distributions, we use discrete Weibull distributions, similar to [11], fitted to the data on steady and non-steady male partners, resulting in the fitted scale and shape parameters in Supplementary Table S1. We explore different truncation/cut-off values for the maximum number of steady and non-steady sexual partners within 12 months, see Supplementary Table S1.

Besides the number of sexual partners, the contact rates are important parameters of the MSM sexual contact network. The contact rates *β_s_* and *β_c_* are defined as the frequency of transmission-relevant sex acts per steady and non-steady sexual partner, respectively. The contact rates *β_s_* and *β_c_* are not directly reported by EMIS-17. Instead, we rely on the reported “Any sex with man” recency data in Figure 5.4 in EMIS-17 to inform the contact rates. Specifically, the contact rates *β_s_* and *β_c_* are estimated by fitting the expected proportions of individuals that has had intercourse within 24 hours, 7 days, 1 month, 6 months and 1 year^6^ (building on the fitted Weibull degree distributions) to the corresponding empirical proportions (given by Figure 5.4 in EMIS-17).

Given the contact rates *β_s_* and *β_c_*, an individual with *d_s_* steady partners and *d_c_* non-steady partners has a total exponential sexual contact rate *β_s_d_s_* + *β_c_d_c_*. Hence, the number of sexual interactions that this individual has in a *k*-day period follows a Poisson distribution with mean *k*(*β_s_d_s_* + *β_c_d_c_*). Thus, the probability that this individual had sex within a *k*-day period follows as 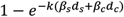, and the expected proportion of individuals that does not have sex during a *k*-day period is

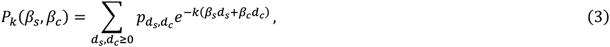

where 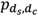 is the probability that an individual has *d_s_* steady partners and *d_c_* non-steady partners, which is specified by the fitted Weibull degree distribution.

We obtain the contact rates *β_s_* and *β_c_* by minimising the Euclidean norm of the expected proportion (3) minus the empirical proportion 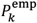 for the time period *k* reported by Figure 5.4 in EMIS-17, i.e.,

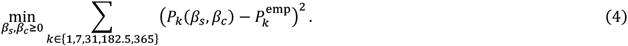

The resulting contact rates are *β_c_* = 3.23**/**365 and *β_s_* = 34.10**/**365. Note that this contact rates are obtained without truncating the degree distribution.

#### S2.3 Model initialisation

The initial state of the model is obtained in four steps. First, to capture the residual population-level protection from historical smallpox vaccination in line with [13], we consider that 14.81% of the MSM population is immune, which is captured by moving these individuals into the vaccinated *V* compartment at time *t* = 0. We choose these individuals uniformly at random.

Second, we consider naturally-acquired immunity from the MPXV clade II outbreak in 2022 by setting the number of individuals in the recovered compartment *R* to either 0, 66, 133, or 266 in a scenario analyses, see Supplementary Table S1. We randomly select initially recovered individuals, with probabilities proportional to their number of sexual contacts (steady plus non-steady degree). This degree-proportional sampling considers that sexually active individuals are more likely to have acquired an MPXV clade II infection than less sexually active individuals.

Third, we consider mpox-specific vaccination campaigns since 2022. Specifically, we additionally move 0%, 5% and 10% (different scenarios) of the 10,000 MSM individuals to the vaccinated compartment *V*. These individuals are chosen uniformly at random from the 15% and 30% (different scenarios) susceptible individuals that remain after steps 1 and 2 with the highest number of sexual partners.

Fourth, the initially infected individual is chosen at random from the remaining susceptible MSM population, by sampling proportionally to the number of the individuals’ sexual contacts (steady plus non-steady degree). This initially infected individual is moved to the undetected infectious compartment *I_N_*.

#### S2.4 Model calibration

Given the initial state obtain as described in A2.4, we calibrate the model to the number of MPXV clade II cases by modifying the contact rates *β_s_* and *β_c_* for each of the 72 MSM communities. Specifically, for each community *l* = 1, …,72, we modify the contact rates *β_s_* and *β_c_* obtained by (4) to *β_s_* ← *m_l_β_s_* and *β_c_* ← *m_l_β_c_*, where the scalar *m_l_* > 0 reflects the relative level of sexual activity in MSM community *l*. While the modified contact rates *β_s_* and *β_c_* do not minimise (4) anymore, the ratio *β_s_***/***β_c_* of contacts per steady versus non-steady sexual partner remains the same.

For each community and parameter setting *l*, the scalar *m_l_* is obtained by minimising the difference between the average final *F*_avg_(*m_l_*) size and the number of MPXV clade II cases after 2022, see Supplementary Table S1. The average final size *F*_avg_(*m_l_*) equals the number of individuals in the compartments *QR_T_* and *QR_D_* at the end of the outbreak, averaged over 5,000 runs of the model. Since the final size *F*_avg_(*m_l_*) is monotonically increasing with respect to the scalar *m_l_*, we employ the bisection method to find the scalar that minimises the difference of the final size *F*_avg_(*m_l_*) to the number of MPXV clade II cases. Across different communities *l*, the fitted scalar ranged from *m_l_* = 0.32 to *m_l_* = 1.64.

### S3. Additional figures and tables

**Supplementary Table S4.**
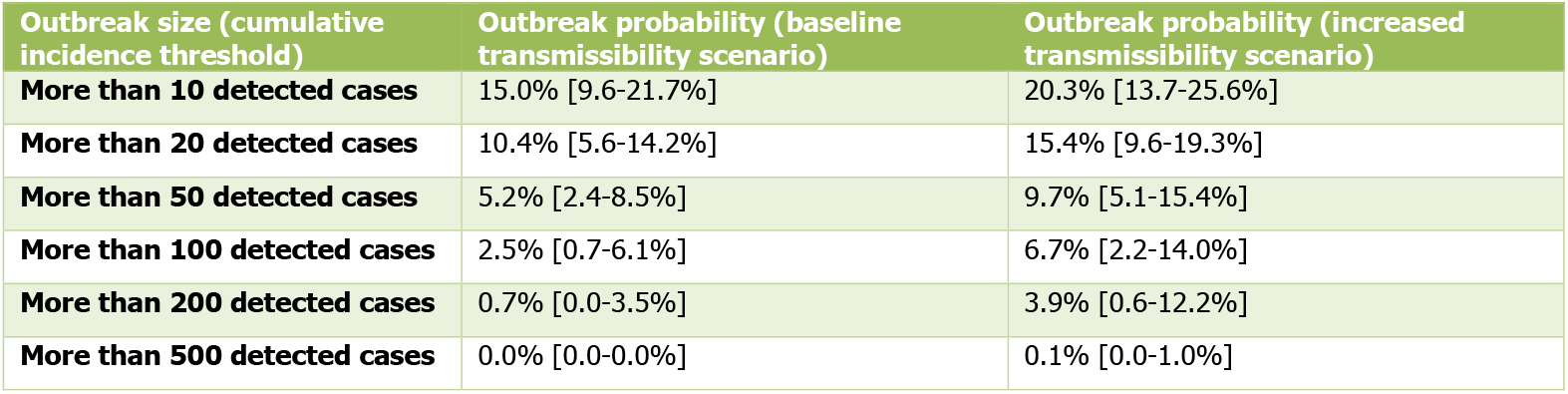
The probability of an MPXV clade Ib outbreak in an MSM population in the EU/EEA starting with a single (undetected) infectious individual for different cumulative incidence thresholds. The baseline transmissibility scenario considers that clade Ib and clade II have the same transmissibility, and the increased transmissibility scenario considers that clade Ib is 15% more transmissible compared to clade II. The table provides the estimated values for the outbreak probabilities (rounded to two digits). The first column shows the outbreak size, and the second and third column show the median and the 90% UI of the outbreak probability for the baseline and increased transmissibility scenarios, respectively.

**Supplementary Table S5.** The effect of increasing the vaccine uptake on reducing the probability of an MPXV clade Ib outbreak exceeding 10 or 100 cases in an MSM population in the EU/EEA starting with a single (undetected) infectious individual. Different rows correspond to different scenarios for the vaccine uptake and vaccine target group. Each entry in the last three columns captures the uncertainty due to different characteristics of the MSM population, including variations in the sexual contact patterns and different immunity levels due to current mpox vaccine uptake (as of November 2024) and past clade II infections. Additionally, each entry includes both clade Ib transmissibility scenarios. The table provides the estimated median and 90% UI of the outbreak probabilities (rounded to two digits).

| Increase of the vaccine uptake | Vaccine targeting level | Probability outbreaks over 10 detected cases | Probability outbreaks over 100 detected cases |
| --- | --- | --- | --- |
| Baseline | N/A | 19.1% [11.2-26.4%] | 4.0% [0.7-13.2%] |
| Low increase of vaccination uptake | Low | 17.4% [9.6-24.3%] | 2.8% [0.4-9.4%] |
|  | Medium | 15.9% [8.0-22.7%] | 1.8% [0.2-7.5%] |
|  | High | 14.7% [7.1-21.6%] | 1.0% [0.1-5.2%] |
| High increase of vaccination uptake | Low | 15.5% [7.8-22.6%] | 1.6% [0.2-6.8%] |
|  | Medium | 12.8% [6.4-19.7%] | 0.1% [0.0-3.3%] |
|  | High | 10.3% [4.4-16.6%] | 0.1% [0.0-1.1%] |

**Supplementary Figure S2.**
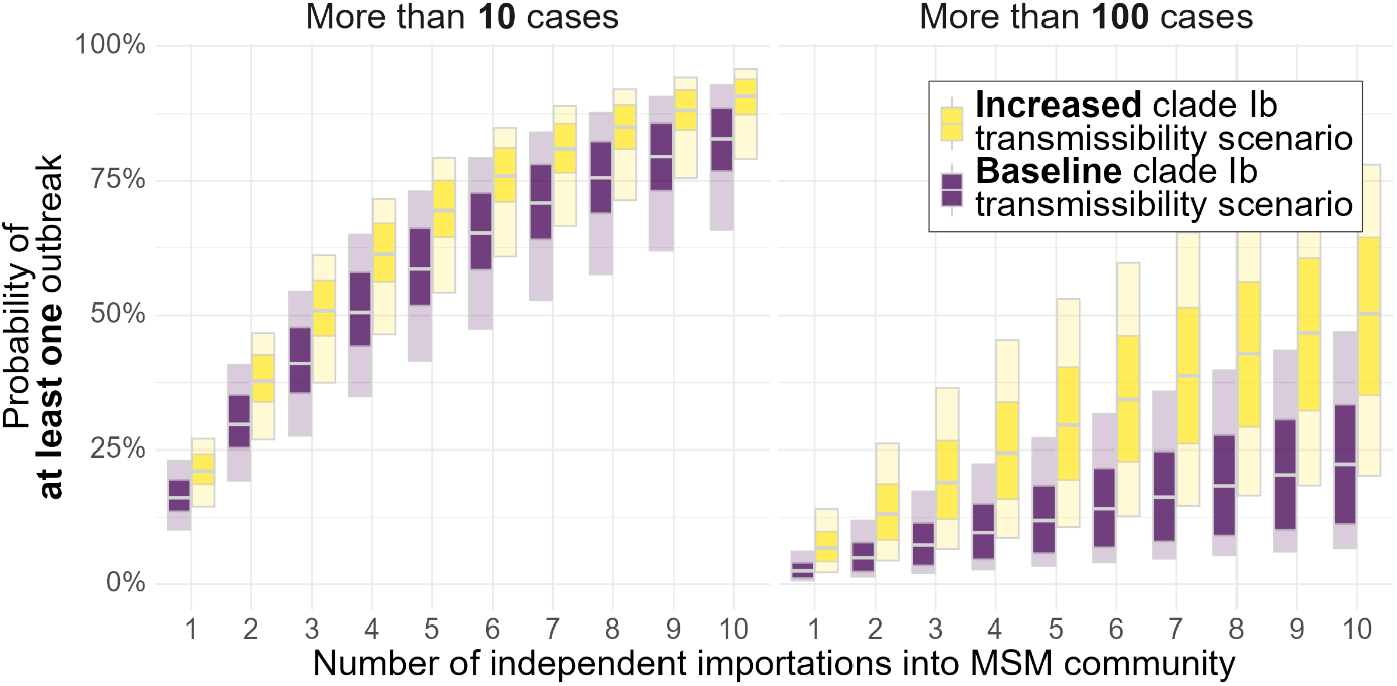
The probability of at least one MPXV clade Ib outbreak exceeding 10 or 100 cases, respectively, in an MSM population in the EU/EEA starting with a single (undetected) infectious individual versus the number of independent importations of MPXV clade Ib. The baseline transmissibility scenario considers that clade Ib and clade II have the same transmissibility, and the increased transmissibility scenario considers that clade Ib is 15% more transmissible compared to clade II. The bars capture the uncertainty due to different characteristics of the MSM population, including variations in the sexual contact patterns and different immunity levels due to mpox vaccine uptake and past clade II infections. The dark and light bars correspond to the 50% and 90% uncertainty interval (UI), respectively, and the grey line shows the median.

## Footnotes

1 While it is difficult to accurately estimate the *absolute* probabilities of importations, estimating the *relative* importation risk is more amenable. For instance, the EpiRisk tool [18] builds upon data on the airline transportation network.

2 For instance, Dublin, Brussels, Copenhagen, Milan, and Prague all have a population size between 1.0-1.3 million. Assuming that approximately 50% of the population are men, 60-70% are in a sexually active age, and approximately 2.8% of men are MSM [19] results in an approximate range of 8,400-12,740 MSM individuals in these cities.

3 In contrast, one could select the initial infection uniformly at random, which would result in a smaller outbreak probability. However, we consider uniform seeding to be less plausible than preferential seeding.

4 As suggested by Schmidt AJ (LSHTM). Personal communication, 8 August 2024.

5 Since the baseline transmissibility scenario assumes the same transmissibility of both clades, the results of this scenario also apply to the introduction of clade II to an MSM community in which there was no large outbreak past 2022.

6 We exclude reported sex recency beyond the preceding 12 months (“5 yrs” and “over 5 yrs” in Figure 5.4 in EMIS-17) from the analysis, as the network structure is assumed to have undergone substantial changes over a five-year period, particularly regarding non-steady partners.

